# TREATMENT PROFILES AND CLINICAL OUTCOMES OF COVID-19 PATIENTS AT PRIVATE HOSPITAL IN JAKARTA

**DOI:** 10.1101/2020.10.14.20212449

**Authors:** Diana Laila Ramatillah, Suri Isnaini

**Affiliations:** Faculty of Pharmacy, Universitas 17 Agustus 1945 Jakarta

**Keywords:** COVID-19, Treatment Profile, Clinical Outcome, Survival Analysis

## Abstract

**Background:** Severe Acute Respiratory Syndrome Coronavirus-2 (SARS-CoV-2) is a virus that causes COVID-19, which has become a worldwide pandemic. However, until now, there is no vaccine or specific drug to prevent or treat COVID-19.

**Objectives:** To find out the effective treatment as an antiviral agent for COVID-19, to determine the correlation between sociodemography with clinical outcomes and duration of treatment, and to determine the relationship between comorbidities with clinical outcomes and duration of treatment for COVID-19 patients.

**Methods:** A prospective cohort study was conducted in this study. This study included only confirmed COVID-19 patients who were admitted to the hospital during April-May 2020. Convenience sampling was used to select 103 patients, but only 72 patients were suitable for inclusion.

**Results:** The survival analysis for COVID-19 patients using the Kaplan Meier method showed that patients receiving Oseltamivir + Hydroxychloroquine had an average survival rate of about 83% after undergoing treatment for about ten days. Gender (p = 0.450) and age (p = 0.226) did not have a significant correlation with the duration of treatment for COVID-19 patients. Gender (p = 0.174) and age (p = 0.065) also did not have a significant correlation with clinical outcome of COVID-19 patients. Comorbidities showed a significant correlation with duration of treatment (p = 0.002) and clinical outcome (p = 0.014) of COVID-19 patients.

**Conclusion:** The most effective antiviral agent in this study based on treatment duration was the combination of Oseltamivir + Hydroxychloroquine. The higher the patient’s average treatment duration, the lower the average survival rate for COVID-19 patients.

## INTRODUCTION

Coronaviruses (CoV) is a single-stranded RNA virus that belongs to the Coronaviridae family. They spread between multiple hosts, clinically presenting with various symptoms, from flu-like to severe, sometimes deadly respiratory infections (Drożdżal *et al*., 2020). The new virus, which is responsible for this pandemic, was initially referred to as “2019-nCoV” but has been renamed “SARS-CoV-2” by the Coronavirus Study Group (CSG). This body is part of the International Committee on Taxonomy of Viruses. (ICTV), It is believed to be familiar with SARS CoV, the pathogen that causes Severe Acute Respiratory Syndrome (SARS). SARS-CoV-2 was recently closely related to SARS-CoV, sharing 80% identity in the RNA sequence (Gorbale *et al*., 2020; Chan *et al*., 2020).

With the first case in humans recorded in December 2019, SARS-CoV-2 is responsible for an outbreak of a respiratory disease called COVID-19 (Coronavirus Diseases 2019). The full spectrum of COVID-19 ranges from mild self-limiting respiratory disorders to severe progressive pneumonia, multiple organ failure, and death (Huang *et al*., 2020). In the central Chinese province of Hubei, Wuhan’s city has been declared the pandemic epicenter, with the Huanan seafood market being one of the first locations where SARS-CoV-2 could potentially cross the species barrier at the animal-human interface. Initial research conducted in Shenzhen, by a group of doctors and scientists from the University of Hong Kong, provided the first evidence that SARS-CoV-2 can be transmitted from human to human (Chan *et al*., 2020).

The Chinese Centers for Disease Control and Prevention reported in a study of 44,672 people (1,023 deaths) that cardiovascular disease, hypertension, diabetes, respiratory disease, and cancer were associated with an increased risk of death (Deng *et al*., 2020). Advanced age, the presence of comorbidities, especially hypertension, diabetes, obesity, and smoking, are factors that increase the risk of severe disease presentation (Lippi *et al*., 2020). However, the characteristics of COVID-19 may differ depending on the demographic and epidemiological profile of each country (Drożdżal *et al*., 2020). In Indonesia, 51.9% of confirmed cases are men, 31.4% of confirmed cases are aged between 31-45 years, with the highest percentage of deaths between 46-59 years of age at 39.4% and 50.5% of confirmed cases of having hypertension as a comorbid disease (Task Force COVID-19, 2020). Assessing the instantaneous mortality rate at any time during follow-up for specific risk factors is essential for determining the suitability of mitigation strategies and for setting priorities for controlling the COVID-19 pandemic. This is especially the case in countries that are currently focusing their efforts on tackling a pandemic (Drożdżal *et al*., 2020).

The COVID-19 pandemic requires the rapid development of a useful therapeutic strategy, in which three concepts are applied: (i) The first approach relies on testing is currently known antiviral agents and verifying their clinical utility (Kim *et al*., 2016; Lu, 2020). (ii) Other modalities are based on molecular libraries and databases, enabling high computing power and simultaneous verification of millions of potential agents (Lu, 2020; Channappanavar *et al*., 2017). (iii) The third strategy involves targeted therapy, which is intended to disrupt the viral genome and function. Properly particles designed to interfere with important steps of viral infection, such as binding to cell surfaces and internalization. Unfortunately, in vitro activity does not necessarily mean success in vivo testing, due to different pharmacodynamic and pharmacokinetic properties (Lu, 2020; Zumla *et al*., 2016). The main group of therapeutic agents that can be useful in the treatment of COVID-19 involves antiviral drugs, certain antibiotics, antimalarials, and immunotherapy drugs (Drożdżal *et al*., 2020).

## METHODS

### Research Design

This research is descriptive, a study conducted to provide an accurate description or description of a situation. The study design used a cohort design. A cohort study was undertaken to support an association between suspected cause and disease, which was measured prospectively. The reason researchers use prospective measurements is to get real data and can avoid bias. The type of data collected is primary data in the form of medical records.

### Samples

Samples used in this study were patients with confirmed COVID-19 who were undergoing treatment and receiving antiviral agent therapy. The inclusion and exclusion criteria in this study are:

Inclusion criteria:

a. Patients demonstrated positive for COVID-19 are undergoing treatment at the hospital.
b. Patients receiving favipiravir and / or Oseltamivir and / or Chloroquine and / or Hydroxychloroquine at the hospital.

Exclusion criteria:

a. Patients confirmed positive for COVID-19 with comorbidities HIV / AIDS.
b. The patient confirmed positive for COVID-19 with a cancer-related disease.
c. The patient tested positive for COVID-19 with pregnancy.

### Data Retrieval Method

Data were taken using a *convenience sampling technique*, namely sampling based on the availability of elements and the ease of obtaining them. Samples were taken or selected because the models were at the right place and time; in this case, all existing samples matched the inclusion during the one month in the hospital.

### Tools and Materials

1. Medical Records Files containing notes and information regarding the patient’s identity, examination data, medication, actions, and other services that have been provided to patients.
2. Observation Sheet Used to load data to be recorded from the patient medical record. The data contains the patient’s identity, medical history, medical history, and clinical status.
3. Ethical Clearance Ethical Eligibility Letter issued by the Faculty of Health, Esa Unggul University with Number: 0303-20.283/DPKE-KEP/FINAL-EA/UEU/IX/2020

## RESULTS AND DISCUSSION

### Effect of Sociodemography on Duration of Treatment for COVID-19 Patients

Based on **Table 1**. it can be seen that based on gender, most of the patients were male, as many as 45 people (62.5%). Data from the Task Force for the Acceleration of Handling COVID-19 in Indonesia as of August 6, 2020, the number of male patients with confirmed COVID-19 was 52.1% and women 47.9% (COVID-19 Task Force, 2020). There is no consistent pattern in who is more likely to be diagnosed with COVID-19. A confirmed diagnosis means that laboratory tests have been carried out. In other words, globally, there is almost the same number of cases between men and women. There is no evidence from this national survey data that men are more likely to contract it than women (Global Health 50/50, 2020).

**Table 1.** Effect of Sociodemography on Duration of Treatment for COVID-19 Patients.

| Indicators | Duration of Treatment |  | Total | Sig.<br>(*P*-<br>Value) |
| --- | --- | --- | --- | --- |
|  | ≤ 14 days<br>n (%) | > 14 days<br>n (%) |  |  |
| **Gender** |  |  |  |  |
| a. Male | 30 (60,0) | 15 (40,0) | 45 | 0,450 |
| b. Female | 22 (55,5) | 12 (45,5) | 27 |  |
| **Age** |  |  |  |  |
| a. 19 - 38 years | 6 (37,5) | 10 (62,5) | 16 | 0,226 |
| b. 39 - 58 years | 22 (68,8) | 10 (31,2) | 32 |  |
| c. 59 - 78 years | 13 (59,1) | 9 (40,9) | 22 |  |
| d. 79 - 85 years | 1 (50,0) | 1 (50,0) | 2 |  |

*Chi-Square Test*

Based on age, most patients were in the 39-58 years age group, as many as 32 people (44.4%), and the least number in the 79-85 years age group was 2 people (2.8%). Data from the Task Force for the Acceleration of Handling COVID-19 in Indonesia as of August 6 2020, the highest number of confirmed COVID-19 patients was in the 31-45 year age group as much as 31.4% (COVID-19 Task Force, 2020). In the 2020 Chinese study, the mean age of COVID-19 patients was 56 years, ranged from 18-87 years, and most patients were male (Zhou *et al*., 2020). Whereas in the study of Guan *et al*., the patients’ mean age was 47 years, and 41.9% of patients were women (Guan *et al*., 2020).

Sig value. Gender and age showed a value of> 0.05, which means that sex and age had no effect on the length of treatment for COVID-19 patients. There are still few studies investigating the duration of hospitalized COVID-19 patients during the pandemic. The mean duration of stay due to COVID-19 has been reported in several studies in China as 10-13 days (Guan *et al*., 2020; Wang *et al*., 2020). However, the length of stay depends on various factors, such as the time elapsed from exposure to the onset of symptoms, and from the time of onset to the time of hospital admission, as well as various factors related to the country-specific context (Thai *et al*., 2020).

### Effect of Sociodemography on *Clinical Outcome of* COVID-19 Patients

Based on **Table 2**. above, it is known that of the 45 male patients, most of them get *clinical outcomes*, as many as 30 people (66, 7%), and 15 people (33.3%) died. Likewise, most of the 27 people with female patients also received *clinical outcomes*, namely as many as 22 people (81.5%) and 5 people (18.5%) died. Gender has no influence on the *clinical outcome* of COVID-19 patients with P-value 0.174> 0.05.

**Table 2.** Effect of Sociodemography on *Clinical Outcome of* COVID-19 Patients.

| Indicators | Clinical Outcome |  | Total | Sig.<br>(P-Value) |
| --- | --- | --- | --- | --- |
|  | Healed<br>n (%) | Death<br>n (%) |  |  |
| Gender |  |  |  |  |
| a. Male | 30 (66,7) | 15 (33,3) | 45 | 0,174 |
| b. Female | 22 (81,5) | 5 (18,5) | 27 |  |
| Age |  |  |  |  |
| a. 19 - 38 years | 14 (87,5) | 2 (12,5) | 16 | 0,065 |
| b. 39 - 58 years | 23 (71,8) | 9 (28,2) | 32 |  |
| c. 59 - 78 years | 15 (68,2) | 7 (31,8) | 22 |  |
| d. 79 - 85 years | 0 (0,0) | 2 (100,0) | 2 |  |
| Chi Square Test |  |  |  |  |

Furthermore, it is known that out of 16 patients aged 19 - 38 years, most of them received *clinical outcomes*, namely 14 people (87.5%), then from 32 patients aged 39 - 58 years, most of them received *clinical outcomes*, which were 23 people (71, 8%). Most of the patients aged 59-78 years received *clinical outcomes*, as many as 15 people (59.1%) and the two patients aged 79-85 all (100%) died. S value. Age has no influence on the *clinical outcome* of COVID-19 patients with P-value 0.065> 0.05. So it can be concluded that sociodemography does not affect the clinical outcome of COVID-19 patients.

Older age, higher SOFA scores, and d-dimers of more than 1 μg / mL are associated with an increased likelihood of death (Zhou *et al*., 2020). In Chenstudy *et al.’s*, the number of patients infected with COVID-19 was more male than female (Chen *et al*., 2020). MERS-CoV and SARS-CoV were also found to infect more men than women (Badawi *et al*., 2016 and Channappanavar *et al*., 2017). The reduction in women’s susceptibility to viral infections can be attributed to the protection of the X chromosome and sexual hormones, which play an essential role in innate and adaptive immunity (Jaillon *et al*., 2019). Chen *et al*. showed that SARS-CoV-2 was more likely to infect older adult men with chronic comorbidities due to the weaker immune function of these patients (Chen *et al*., 2020).

### Effect of Comorbidities on Treatment Duration of Covid-19 Patients

Based on **Table 3**., it is known that of the seven patients who did not have comorbidities, most of them underwent treatment ≤ 14 days, as many as six people (85.7%). Another case with patients with comorbid pneumonia is that most of the ten people also underwent treatment duration of more than14 days, as many as nine people (90.0%), while patients who had comorbid pneumonia - degenerative diseases mostly underwent treatment duration, less than 14 days as many as 35 people (63.6%). Comorbidities have a relationship with the treatment length for COVID-19 patients with a P-value of 0.002 <0.05. Based on the study of Wang *et al*., The median duration from first symptoms to dyspnea, hospital admission, and ARDS was 5 days, 7 days and 8 days, respectively (Wang *et al*., 2020).

**Table 3.** Effect of Comorbidities on Treatment Duration of Covid-19 Patients.

| Comorbidities | Duration of Treatment |  | Total | Sig. (P-Value) |
| --- | --- | --- | --- | --- |
|  | ≤ 14 days | > 14 days |  |  |
|  | n (%) | n (%) |  |  |
| a. Without Commorbid | 6 (85,7) | 1 (14,3) | 7 | 0,002 |
| b. Pneumonia | 1 (10,0) | 9 (90,0) | 10 |  |
| c. Pneumonia – Degenerative diseases | 35 (63,6) | 20 (36,4) | 55 |  |

*Chi Square Test*

### Effect of Commorbidities on Clinical Outcomes of Covid-19 Patients

Based on **Table 4**. it is known that of the 7 patients who did not have comorbidities, all (100.0%) had *clinical outcomes* cured. Likewise with patients who had comorbidities with pneumonia, out of 10 people (100.0%) had a *clinical outcome* cured.patients who had comorbidities with most of pneumonia - degenerative diseases received *clinical outcomes*, as many as 35 people (63.6%) and the remaining 20 people (36.4%) died. Sig value shows a value of 0.014 <0.05, which means that comorbidities have a relationship with the *clinical outcome* of COVID-19 patients.

**Table 4.**
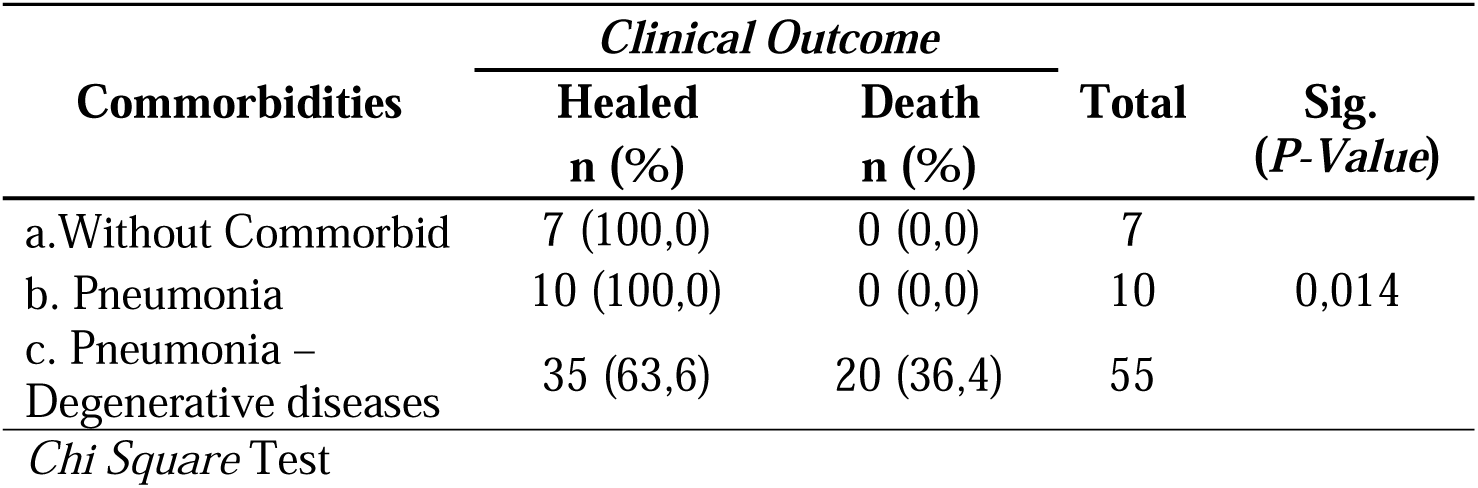
Effect of Commorbidities on Clinical Outcomes of Covid-19 Patients.

| Commorbidities | Clinical Outcome |  | Total | Sig. (P-Value) |
| --- | --- | --- | --- | --- |
|  | Healed | Death |  |  |
|  | n (%) | n (%) |  |  |
| a. Without Commorbid | 7 (100,0) | 0 (0,0) | 7 | 0,014 |
| b. Pneumonia | 10 (100,0) | 0 (0,0) | 10 |  |
| c. Pneumonia – Degenerative diseases | 35 (63,6) | 20 (36,4) | 55 |  |

*Chi Square Test*

Several reports suggest that COVID-19 can cause severe, sometimes fatal pneumonia (Wang *et al*., 2020 and Chen *et al*., 2020). Besides, about half of the patients infected with SARS-CoV-2 had a chronic underlying disease, particularly cardiovascular and cerebrovascular disease, and diabetes, similar to MERS-CoV (Badawi *et al*., 2020). The risk of death at any time during follow-up was significantly higher in men, individuals in the older age group, with chronic kidney disease, and people who were hospitalized (Salinas-Escudero *et al*., 2020).

The Chinese Centers for Disease Control and Prevention reported in a study of 44,672 people (1,023 deaths) that cardiovascular disease, hypertension, diabetes, respiratory disease, and cancer were associated with an increased risk of death (Deng *et al*., 2020). However, correction for association with age is not possible (Williamson *et al*., 2020). A cross-sectional survey in the UK of 16,749 patients hospitalized with COVID-19 showed that the risk of death was higher in patients with heart, lung, and kidney disease, as well as cancer, dementia and obesity (Docherty *et al*., 2020).

### Overview of the Effectiveness of COVID-19 Antiviral Agents

Based on **Table 5**. it can be seen that from 16 patients who received Oseltamivir therapy Most of them got clinical outcomes cured, namely 13 people (81.3%). From 28 patients who received combination therapy of Oseltamivir + Chloroquine, 16 people (57.1%) got clinical outcome*s*. Of the patients who received combination therapy of Oseltamivir + Hydroxychloroquine, 4 (50%) received clinical outcomes, and 4 (50%) died. Of the 8 patients who received the combination of Favipiravir + Chloroquine therapy, all (100%) had a clinical outcome. Most of the patients who received combination therapy Favipiravir + Oseltamivir + Chloroquine received clinical outcomes, as many as 11 people (91.67%). Sig value shows a value of 0.025 <0.05, which means that COVID-19 Antiviral Agent Therapy has an influence on the clinical outcome of COVID-19 patients.

**Table 5.** Overview of the Effectiveness of COVID-19 Antiviral Agents.

| Regimen | Clinical Outcome |  | Total | Sig. (P-Value) |
| --- | --- | --- | --- | --- |
|  | Healed<br>n (%) | Death<br>n (%) |  |  |
| Oseltamivir | 13 (81,3) | 3 (18,7) | 16 | 0,025 |
| Oseltamivir + Klorokuin | 16 (57,1) | 12 (42,9) | 28 |  |
| Oseltamivir + Hidroksiklorokuin | 4 (50,0) | 4 (50,0) | 8 |  |
| Favipiravir + Klorokuin | 8 (100,0) | 0 (0,0) | 8 |  |
| Favipiravir + Oseltamivir + Klorokuin | 11 (91,67) | 1 (8,33) | 12 |  |

The Indonesian Lung Doctors Association recommends Oseltamivir for the treatment of COVID-19 because the drug is easily accessible in Indonesia and has been produced domestically (Rossa, 2020). In a previous study, a combination of low-dose favipiravir with Oseltamivir showed a synergistic response to influenza virus infection in white mice (Smee et al., 2010). The results of a study led by the China Pneumonia Research Network prove that favipiravir combined with Oseltamivir is better than Oseltamivir alone in the treatment of severe influenza (Wang et al., 2019). Oseltamivir has also been used in clinical trials in various combinations with Chloroquine and favipiravir, a nucleoside analogue known as a broad-spectrum antiviral drug that has shown EC50 61.88 μM against SARS-CoV-2 and low toxicity (CC50> 400 μM) (Cao et al. ., 2020).

To date, no specific treatment has been recommended for coronavirus infection except for careful supportive care (de Wit et al., 2016). The approach taken for this disease is to control the source of infection, use of personal protective precautions to reduce the risk of transmission, and early diagnosis, isolation, and supportive care for infected patients. Antibacterial agents are not sufficient. Besides, there are no antiviral agents that are useful for treating SARS and MERS (Wang et al., 2020). All patients in the study received antibacterial agents, 90% received antiviral therapy, and 45% received methylprednisolone. The doses of Oseltamivir and methylprednisolone vary depending on the severity of the disease. However, no significant results were observed (Wang et al., 2020).

Chloroquine (CQ) and its hydroxyl analog, Hydroxychloroquine (HCQ), are weak bases used as antimalarial agents for half a century. Apart from these antimalarial activities, Chloroquine and Hydroxychloroquine have gained interest in other areas of infectious diseases. For viruses, Chloroquine causes coating inhibition and/or changes in post-translational modification of newly synthesized proteins, especially inhibition of glycosylation (Rolain *et al*., 2020). Chloroquine or Hydroxychloroquine may also have indirect antiviral effects. Chloroquine was found to be effective in preventing the spread of coronavirus (CoV) associated with the severe acute respiratory syndrome (SARS) in cell culture by interfering with terminal glycosylation of the cellular receptor, angiotensin-converting enzyme 2 (ACE2) (Vincent *et al*., 2005); and sialic acid, whose biosynthesis can be inhibited by Chloroquine or Hydroxychloroquine, is a component of the SARS-CoV receptor and orthomyxovirus (Savarino *et al*., 2006).

Research in China studied Chloroquine’s effects in vitro, using Vero E6 cells infected with SARS-CoV-2 at a multiplicity of infection (MOI) 0.05. This study shows that Chloroquine is very effective in reducing viral replication, with an Effective Concentration (EC) of 90 of 6.90 μM, which can be easily achieved with standard doses, due to its good penetration in tissues, including in the lungs. Chloroquine is known to block viral infection by increasing endosomal pH and by interfering with the glycosylation of SARS-CoV cellular receptors. The authors also speculate that a known immunomodulatory effect of the drug might enhance the in vivo antiviral effect (Wang *et al*., 2020).

Oseltamivir is a neuraminidase inhibitor approved for influenza treatment; there is no documented in vitro activity against SARS-CoV-2 (Sanders *et al*., 2020). The COVID-19 outbreak in China initially occurred during the peak influenza season, so most patients received empiric oseltamivir therapy until SARS-CoV-2 discovered the cause of COVID-19 (Wang *et al*., 2020). Several clinical trials are currently including Oseltamivir in the comparison arm but not as a proposed therapeutic intervention (*ClinicalTrialsgov*, 2020).

Favipiravir, formerly known as T-705, is a drug from the purine nucleotide group, favipiravir ribofuranosyl-5′-triphosphate. The active agent inhibits RNA polymerase, stopping viral replication. Most of the favipiravir’s preclinical data come from influenza and Ebola activity. However, these agents have also demonstrated broad activity against other RNA viruses (Furuta *et al*., 2017). In vitro, EC_50_ favipiravir against SARS-CoV-2 was 61.88 μM/L in Vero E6 cells (Cao *et al*., 2020). Favipiravir first enters infected cells through endocytosis and is then converted to active ribofuranosyl phosphate favipiravir through phosphoryibosylation and phosphorylation (Furuta *et al*., 2017). Antiviral activity selectively targets the conservative catalytic domain of *RNA-dependent RNA-polymerase* (RdRp) which can inhibit RNA polymerization activity, disrupting the nucleotide incorporation process of Curr Pharmacol Rep during viral RNA replication (Furuta *et al*., 2017). Dysregulation in viral RNA replication results in an increase in the number and frequency of transition mutations, including replacement of guanine (G) by adenine (A) and cytosine (C) by thymine (T) or C by Uracil (U), which induces lethal mutagenesis in viral RNA (Furuta *et al. al*., 2017). Randomized controlled trials have shown that COVID-19 patients receiving favipiravir therapy had a higher recovery rate (71.43%) than those treated with umifenovir (55.86%), and significantly more duration of fever and cough relief,shorter in the favipiravir group than in the umifenovir group (Chen *et al*., 2020).

### *Survival Analysis* Among COVID-19 Patients

Based on **Figure 1**. above, it can be seen that patients who received combination therapy of Oseltamivir + Hydroxychloroquine had an average survival rate of about 83% after undergoing treatment for about ten days. Patients receiving Oseltamivir therapy had an average survival rate of about 18% after about 27 days of treatment. Patients receiving the combination Oseltamivir + Chloroquine therapy had an average survival rate of about 17% after about 23 days of treatment. Patients who received combination therapy Favipiravir + Chloroquine had an average survival rate of about 17% after about 31 days of treatment. Meanwhile, patients who received combination therapy Favipiravir + Oseltamivir + Chloroquine had an average survival rate of about 10% after undergoing treatment for about 39 days.

**Figure 1.**
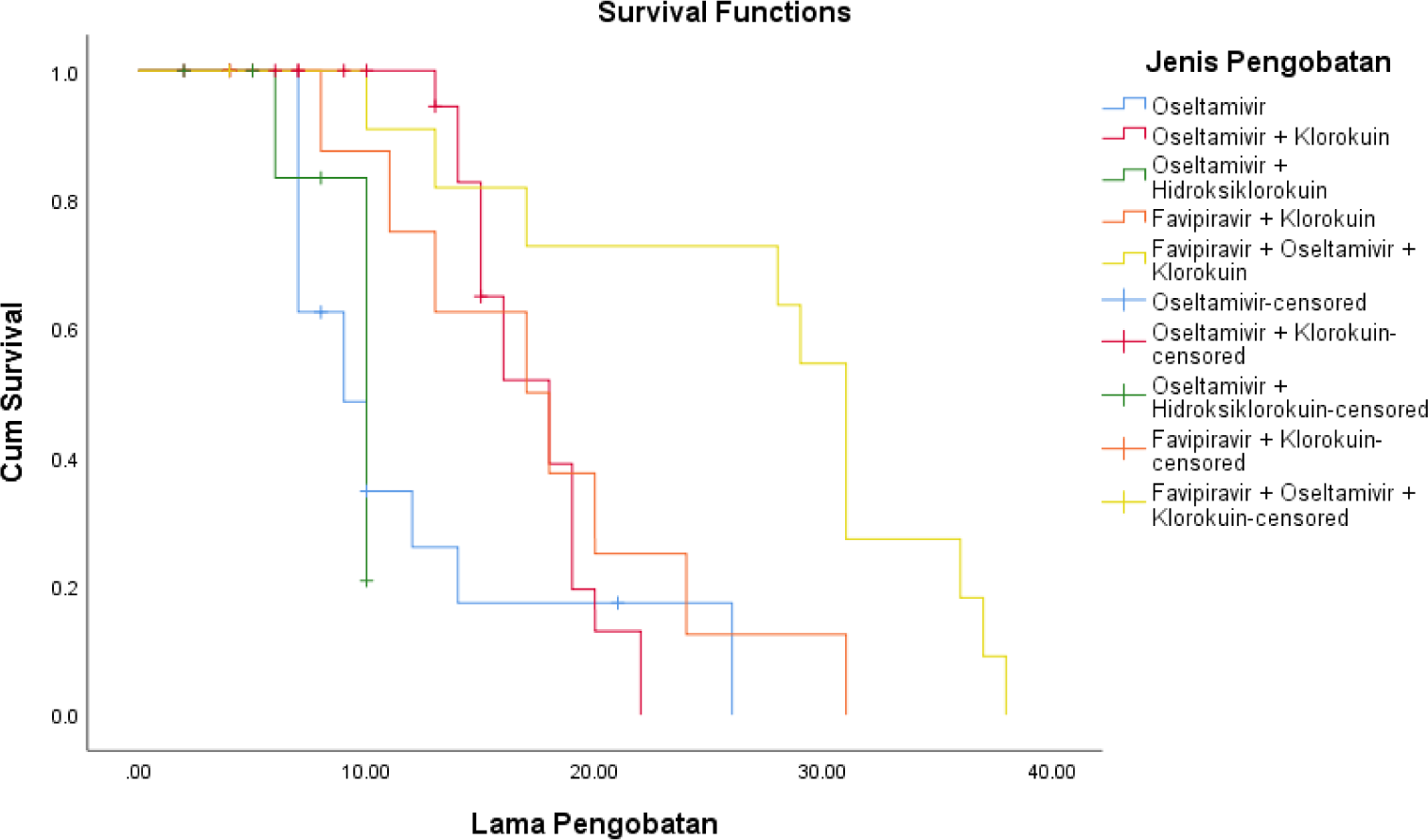
*Survival Analysis* with Kaplan Meier Methods.

A significant association was found among the overall comparisons between the analysis of survival and length of hospitalization of COVID-19 patients. In a study conducted by Thai *et al*., It was shown that the mean duration of hospital stay was 21 (IQR: 16-34) days. The multivariable Cox regression model shows that age, place of residence, and contamination source are significantly associated with longer duration of stay (Thai *et al*., 2020).

In Wang, Z, *et al*., The mean length of stay in the hospital was 19 days (IQR: 14-23, range: 3-41). Adjusted multivariate analysis showed that longer length of stay was associated with a factor of age 45 and over; who is admitted to a provincial hospital; and those who are seriously ill. There is no gender difference (Wang, Z, *et al*., 2020).

## CONCLUSION

The most widely used antiviral agent therapy for patients with confirmed COVID-19 was the combination of Oseltamivir + Chloroquine in 28 patients (38.9%). Based on the Chi-Square test, it was found that there was a significant relationship between COVID-19 antiviral agent therapy and the clinical outcome of COVID-19 patients (p = 0.025). The antiviral agent therapy that was rated the most effective based on the length of treatment was the combination of Oseltamivir + Hydroxychloroquine which had the highest survival rate at around 83% after undergoing treatment for about 10 days (p = 0.027). The number of COVID-19 patients who died was 20 people (27.8%), while 52 people recovered (72.2%).

Based on gender, most of the patients were male, as many as 45 people (62.5%), and by age, most patients were in the 39-58 years age group as many as 32 people (44.4%). In the Chi-Square test, it was found that there was no significant effect between gender (p = 0.450) and age (p = 0.226) on the length of treatment for COVID-19 patients. Based on the Chi-Square test, there was no significant effect between gender (p = 0.174) and age (p = 0.065) on the clinical outcome of COVID-19 patients.

Most patients had comorbidities with pneumonia-degenerative diseases, as many as 55 people (76.4%). The Chi-Square test showed a significant relationship between the types of comorbidities and the length of treatment for COVID-19 patients (p = 0.002). Based on the Chi-Square test, there was a significant relationship between the types of comorbidities and the clinical outcome of COVID-19 patients (p = 0.014).

## LIMITATION

The limitations that occur in this study are the relatively short research time; the extraordinary volume and speed of published literature on the treatment of COVID-19 means that research findings and recommendations continue to evolve as new evidence emerges; and treatment data published to date have come exclusively from observational data or small clinical trials (none of which had more than 250 patients), which presented a higher risk of bias or inaccuracy concerning large treatment effect sizes.

## Data Availability

All the data are available in manuscript

