## Supplementary material for "TREATMENT PROFILES AND CLINICAL OUTCOMES OF COVID-19 PATIENTS AT PRIVATE HOSPITAL IN JAKARTA": Table1, Table 2, Table 3, Table 4 and Table 5

| | | | | | |
|---|---|---|---|---|---|
| Male | 39 - 58 | <= 14 days | Pneumonia - Degeneratif Disease | Favipiravir + Oseltamivir + Klorokuin | Die |
| Male | 39 - 58 | <= 14 days | Pneumonia - Degeneratif Disease | Oseltamivir + Hidroksiklorokuin | Die |
| Male | 59 - 78 | <= 14 days | Pneumonia - Degeneratif Disease | Oseltamivir + Klorokuin | Die |
| Female | 59 - 78 | > 14 days | Pneumonia - Degeneratif Disease | Oseltamivir + Klorokuin | Swab Negatif |
| Male | 19 - 38 | > 14 days | Pneumonia | Oseltamivir + Klorokuin | Swab Negatif |
| Female | 39 - 58 | <= 14 days | Pneumonia - Degeneratif Disease | Oseltamivir + Klorokuin | Swab Negatif |
| Male | 19 - 38 | > 14 days | Pneumonia | Oseltamivir + Klorokuin | Swab Negatif |
| Male | 59 - 78 | <= 14 days | Pneumonia - Degeneratif Disease | Oseltamivir + Klorokuin | Die |
| Male | 59 - 78 | <= 14 days | Pneumonia - Degeneratif Disease | Oseltamivir + Klorokuin | Die |
| Male | 79 - 98 | > 14 days | Pneumonia - Degeneratif Disease | Oseltamivir + Klorokuin | Die |
| Female | 59 - 78 | <= 14 days | Pneumonia - Degeneratif Disease | Oseltamivir + Klorokuin | Die |
| Male | 39 - 58 | <= 14 days | Pneumonia - Degeneratif Disease | Oseltamivir + Klorokuin | Die |
| Female | 59 - 78 | > 14 days | Pneumonia - Degeneratif Disease | Oseltamivir | Die |
| Female | 79 - 98 | <= 14 days | Pneumonia - Degeneratif Disease | Oseltamivir | Die |
| Female | 39 - 58 | <= 14 days | Pneumonia - Degeneratif Disease | Oseltamivir + Klorokuin | Die |
| Female | 39 - 58 | <= 14 days | Pneumonia - Degeneratif Disease | Oseltamivir + Hidroksiklorokuin | Die |
| Male | 39 - 58 | <= 14 days | Pneumonia - Degeneratif Disease | Oseltamivir + Klorokuin | Die |
| Male | 59 - 78 | <= 14 days | Pneumonia - Degeneratif Disease | Oseltamivir + Klorokuin | Die |
| Female | 59 - 78 | > 14 days | Pneumonia - Degeneratif Disease | Oseltamivir + Klorokuin | Swab Negatif |
| Male | 39 - 58 | <= 14 days | Pneumonia - Degeneratif Disease | Oseltamivir + Klorokuin | Die |
| Male | 59 - 78 | > 14 days | Pneumonia - Degeneratif Disease | Oseltamivir + Klorokuin | Swab Negatif |
| Female | 39 - 58 | <= 14 days | Pneumonia - Degeneratif Disease | Oseltamivir + Hidroksiklorokuin | Swab Negatif |
| Female | 19 - 38 | > 14 days | Pneumonia - Degeneratif Disease | Favipiravir + Oseltamivir + Klorokuin | Swab Negatif |

| | | | | | |
|---|---|---|---|---|---|
| Male | 39 - 58 | <= 14 days | Pneumonia - Degeneratif Disease | Oseltamivir + Hidroksiklorokuin | Die |
| Male | 19 - 38 | <= 14 days | Pneumonia - Degeneratif Disease | Oseltamivir + Klorokuin | Die |
| Female | 39 - 58 | <= 14 days | Pneumonia - Degeneratif Disease | Oseltamivir + Klorokuin | Swab Negatif |
| Male | 39 - 58 | <= 14 days | Pneumonia - Degeneratif Disease | Oseltamivir + Klorokuin | Die |
| Male | 19 - 38 | <= 14 days | Pneumonia - Degeneratif Disease | Oseltamivir | Die |
| Male | 39 - 58 | > 14 days | Pneumonia - Degeneratif Disease | Oseltamivir + Klorokuin | Swab Negatif |
| Female | 39 - 58 | <= 14 days | Pneumonia - Degeneratif Disease | Oseltamivir + Hidroksiklorokuin | Swab Negatif |
| Female | 19 - 38 | > 14 days | Pneumonia | Oseltamivir + Klorokuin | Swab Negatif |
| Male | 39 - 58 | <= 14 days | Pneumonia - Degeneratif Disease | Oseltamivir + Hidroksiklorokuin | Swab Negatif |
| Male | 19 - 38 | > 14 days | Pneumonia - Degeneratif Disease | Oseltamivir + Klorokuin | Swab Negatif |
| Male | 39 - 58 | > 14 days | Pneumonia - Degeneratif Disease | Oseltamivir + Klorokuin | Swab Negatif |
| Male | 59 - 78 | <= 14 days | Pneumonia - Degeneratif Disease | Oseltamivir | Swab Negatif |
| Female | 59 - 78 | > 14 days | Pneumonia - Degeneratif Disease | Oseltamivir | Swab Negatif |
| Male | 39 - 58 | <= 14 days | Pneumonia - Degeneratif Disease | Oseltamivir | Swab Negatif |
| Male | 39 - 58 | <= 14 days | Pneumonia - Degeneratif Disease | Oseltamivir | Swab Negatif |
| Female | 19 - 38 | <= 14 days | Pneumonia - Degeneratif Disease | Oseltamivir | Swab Negatif |
| Male | 59 - 78 | <= 14 days | Pneumonia - Degeneratif Disease | Oseltamivir | Swab Negatif |
| Female | 39 - 58 | <= 14 days | Pneumonia - Degeneratif Disease | Oseltamivir + Hidroksiklorokuin | Swab Negatif |
| Female | 59 - 78 | <= 14 days | Pneumonia - Degeneratif Disease | Oseltamivir | Swab Negatif |
| Female | 39 - 58 | <= 14 days | Pneumonia - Degeneratif Disease | Oseltamivir | Swab Negatif |
| Male | 59 - 78 | <= 14 days | Pneumonia - Degeneratif Disease | Oseltamivir + Hidroksiklorokuin | Die |
| Male | 19 - 38 | <= 14 days | None | Oseltamivir + Klorokuin | Swab Negatif |
| Female | 19 - 38 | > 14 days | Pneumonia | Favipiravir + Oseltamivir + Klorokuin | Swab Negatif |

| | | | | | |
|---|---|---|---|---|---|
| Female | 39 - 58 | > 14 days | Pneumonia | Oseltamivir + Klorokuin | Swab Negatif |
| Male | 39 - 58 | > 14 days | Pneumonia - Degeneratif Disease | Oseltamivir + Klorokuin | Swab Negatif |
| Male | 19 - 38 | > 14 days | Pneumonia - Degeneratif Disease | Oseltamivir + Klorokuin | Swab Negatif |
| Female | 39 - 58 | <= 14 days | None | Favipiravir + Klorokuin | Swab Negatif |
| Male | 59 - 78 | <= 14 days | None | Favipiravir + Oseltamivir + Klorokuin | Swab Negatif |
| Male | 39 - 58 | <= 14 days | None | Favipiravir + Klorokuin | Swab Negatif |
| Female | 39 - 58 | > 14 days | Pneumonia - Degeneratif Disease | Favipiravir + Oseltamivir + Klorokuin | Swab Negatif |
| Male | 39 - 58 | > 14 days | Pneumonia - Degeneratif Disease | Favipiravir + Oseltamivir + Klorokuin | Swab Negatif |
| Female | 19 - 38 | > 14 days | Pneumonia - Degeneratif Disease | Favipiravir + Oseltamivir + Klorokuin | Swab Negatif |
| Male | 59 - 78 | > 14 days | Pneumonia - Degeneratif Disease | Favipiravir + Klorokuin | Swab Negatif |
| Male | 59 - 78 | <= 14 days | Pneumonia - Degeneratif Disease | Favipiravir + Klorokuin | Swab Negatif |
| Male | 39 - 58 | > 14 days | Pneumonia - Degeneratif Disease | Favipiravir + Oseltamivir + Klorokuin | Swab Negatif |
| Female | 19 - 38 | > 14 days | Pneumonia | Favipiravir + Klorokuin | Swab Negatif |
| Male | 59 - 78 | > 14 days | Pneumonia | Favipiravir + Oseltamivir + Klorokuin | Swab Negatif |
| Male | 39 - 58 | > 14 days | Pneumonia - Degeneratif Disease | Favipiravir + Oseltamivir + Klorokuin | Swab Negatif |
| Male | 59 - 78 | > 14 days | Pneumonia - Degeneratif Disease | Favipiravir + Klorokuin | Swab Negatif |
| Male | 59 - 78 | <= 14 days | Pneumonia - Degeneratif Disease | Favipiravir + Oseltamivir + Klorokuin | Swab Negatif |
| Male | 39 - 58 | > 14 days | None | Favipiravir + Klorokuin | Swab Negatif |
| Male | 19 - 38 | > 14 days | Pneumonia | Favipiravir + Oseltamivir + Klorokuin | Swab Negatif |
| Male | 39 - 58 | <= 14 days | None | Oseltamivir | Swab Negatif |
| Male | 19 - 38 | <= 14 days | Pneumonia | Oseltamivir | Swab Negatif |
| Female | 39 - 58 | > 14 days | Pneumonia | Favipiravir + Klorokuin | Swab Negatif |

| | | | | | |
|---|---|---|---|---|---|
| Male | 59 - 78 | > 14 days | Pneumonia - Degeneratif Disease | Oseltamivir + Klorokuin | Swab Negatif |
| Female | 19 - 38 | <= 14 days | None | Oseltamivir | Swab Negatif |
| Female | 39 - 58 | <= 14 days | Pneumonia - Degeneratif Disease | Oseltamivir | Swab Negatif |
| Male | 59 - 78 | <= 14 days | Pneumonia - Degeneratif Disease | Oseltamivir | Swab Negatif |
